# Weighting the weeks that matter: entropic tilting for epidemic forecast evaluation

**DOI:** 10.64898/2026.11.12.26367949

**Authors:** Joseph Mathews, Justin Lessler, Joseph Lemaitre

## Abstract

The performance of infectious disease forecasting models is commonly calculated under the implicit assumption that forecast accuracy is equally important across time. This approach may obscure or under-weight differences in model performance during the epidemic periods that matter most for public-health decisions. Here, we introduce the weighted prequential score (WPS), a forecast evaluation framework that can emphasize epidemic periods of interest. The WPS is a weighted average of scores over time, where weights are constructed through entropic tilting which up-weights priority periods while keeping the WPS as close as possible to the arithmetic average. We demonstrate the WPS on forecasts of incident hospitalizations submitted to the CDC FluSight challenge, focusing on two priority periods: weeks of rapid growth in incidence and weeks near the seasonal peak. Across season-location-horizon pairs, doubling the weight assigned to priority periods changes the top-ranked model in 10%–26% of eligible settings, while the top-ranked model remains unchanged regardless of the amount of weight assigned in 37%-68% of settings. We demonstrate that for suitable weights, the models favored by the WPS yield an average gain in accuracy during priority periods that is at least as large as the average accuracy lost during the remaining weeks. Because weights can be defined using observed outcomes, the WPS is not a proper scoring rule. Hence, we recommend using the WPS as a retrospective evaluation diagnostic. The WPS can guide public-health practitioners in selecting models that perform well during periods most relevant to their decisions, while encouraging forecasting teams to improve accuracy during these critical weeks.

**Author summary:** In the context of public health decisions, the importance of accurate infectious disease forecasts may vary over the course of an epidemic. For example, accurate forecasts of incident hospitalizations around the seasonal peak may be particularly important for hospital staffing decisions. We introduce a forecast scoring rule that allows users to emphasize accuracy during the epidemic phases most relevant to their decisions. We demonstrate our approach on forecasts of influenza hospitalization incidence, focusing on periods of rapid growth and weeks surrounding the seasonal peak. We find our approach leads to model rankings that prioritize accuracy during these periods with minimal losses in performance across the entire epidemic.

## 1 Introduction

Quantitative evaluation of probabilistic forecasts of infectious disease incidence requires a *scoring rule*. In infectious disease modeling, commonly used rules include the weighted interval score (WIS) [1, 2], continuous ranked probability score (CRPS) [3, 4], and logarithmic score [5, 6]. Each of these scoring rules is proper, meaning that the expected score is minimized when the forecaster issues their true beliefs about the data-generating distribution [1, 4, 7]. To compare the performance of forecasting models over multiple forecasts (e.g., a respiratory disease season), a common approach is to report the arithmetic average of the scores for each forecast-observation pair; see [1, 2, 8] for examples. This method provides an interpretable summary of forecast performance and has well-understood theoretical properties [7, 9–12]. This arithmetic average assigns equal weight to each individual forecast-observation pair; however, the value of an accurate forecast may vary over the course of an epidemic. For example, errors during periods of rapid growth or high incidence might carry greater consequences than errors during stable or decreasing phases, as they may lead to underprepared hospitals and delayed responses.

Consider the following example: Fig 1 shows two forecasts for the 2023/2024 influenza season in Michigan submitted to FluSight, a Centers for Disease Control and Prevention (CDC)-run project that aggregates influenza forecasts for U.S. states from multiple teams [13]. Over the entire season, the `UMass-flusion` model ranked 1st by average WIS among all models that submitted more than 50% of the target weeks, but its 90% prediction intervals did not contain most of the observed incident hospitalization counts near the seasonal peak. Conversely, the `UMass-trends_ensemble` model, which ranked 7th, contained the observed count within its 90% prediction intervals for nearly all of these weeks. If we are concerned about public-health or operational decisions that are made when cases are high, the `UMass-trends_ensemble` model might be more useful than the `UMass-flusion` model, despite the latter achieving a lower average WIS. This suggests that a modified scoring criterion could more accurately reflect the utility of a forecast in many contexts.

**Fig 1.**
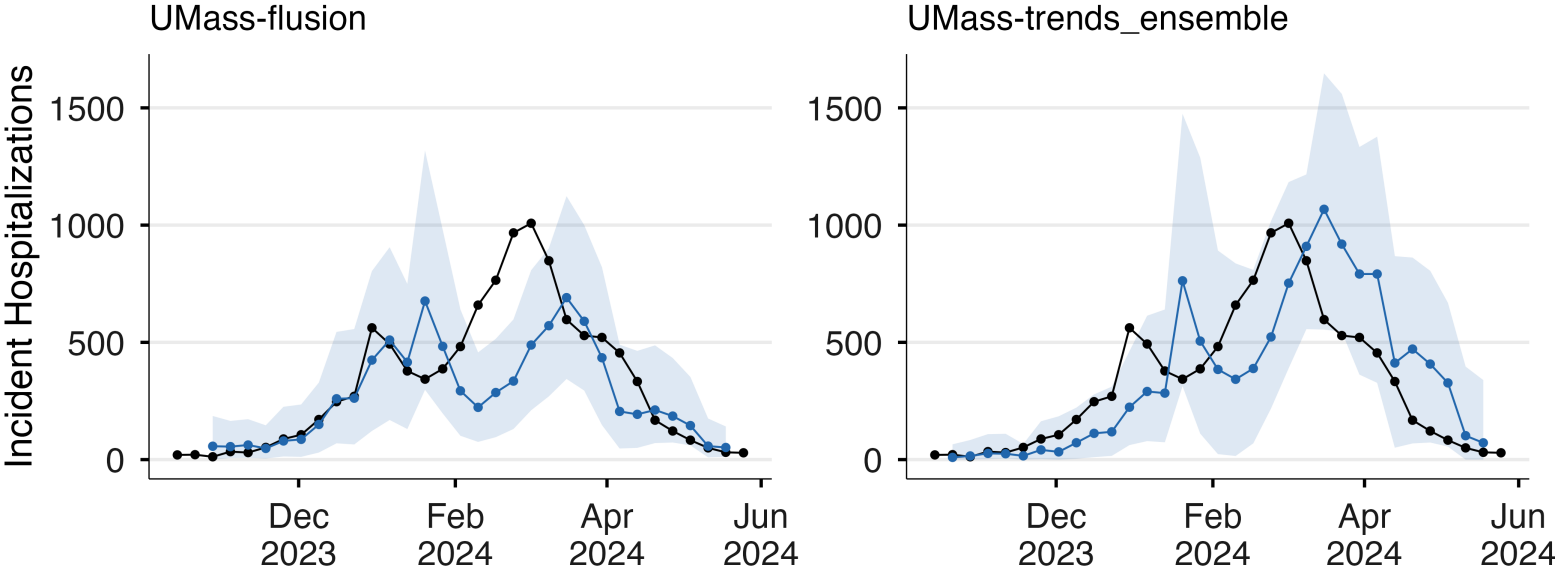
Forecasts of weekly incident influenza hospitalization counts for the state of Michigan at a 2-week horizon for the `UMass-flusion` and `UMass-trends ensemble` models, plotted against the finalized observed value (black lines) for the 2023/2024 season. For a given model, blue lines denote the median forecasts with the shaded region highlighting the 90% prediction intervals.

The adaptation of proper scoring rules to public-health decision making problems has been explored previously [14–16] with approaches that connect forecast evaluation to an explicit definition of utility. However, specifying a utility function can, in many cases, be difficult or impractical. Instead, it may be easier to identify periods when forecast accuracy matters most. These periods of interest, such as weeks surrounding the seasonal peak, may only be identifiable retrospectively. In such cases, a scoring rule emphasizing performance during these periods is generally not proper as it relies on information unavailable at forecast time [17]. Nonetheless, such evaluation methods provide a useful performance diagnostic. In weather forecasting, for example, the U.S. National Weather Service evaluates ensemble forecasts of precipitation conditional on observed precipitation exceeding a chosen threshold [18–20]; similar approaches have also been used to evaluate forecasts for other high-impact weather events [21], including heat waves [22].

Here, we introduce the *weighted prequential score* (WPS), a scoring rule that allows emphasis on forecast performance during designated periods. To do so, the WPS evaluates models using a weighted average of their weekly scores. We construct weights using entropic tilting (ET) [23–25], so that the WPS deviates as little as possible from the arithmetic average. We illustrate the WPS using influenza hospitalization forecasts submitted to the CDC FluSight Forecast Hub during the 2023/2024, 2024/2025, and 2025/2026 seasons. We consider two illustrative priority periods: weeks surrounding the seasonal peak and high-growth weeks. We explore the sensitivity of the rankings of FluSight models to the emphasis ratio *r*, defined as the weight assigned to an event week relative to a non-event week, and how the contribution of weekly scores to the WPS changes under different priorities. Finally, we revisit the *forecaster’s dilemma* [17] and discuss its implications for our approach.

## 2 Weighted forecast evaluation framework

In this section, we introduce the weighted prequential score (WPS) and derived quantities that summarize its effect on model ranking. For simplicity, we describe our method assuming single-week-ahead forecasts, although it generalizes to any forecast horizon.

### 2.1 Weighted prequential score

Let *Y*_1:*T*_ = (*Y*_1_, … , *Y*_*T*_) be a sequence of weekly observations over a season of duration *T* weeks (e.g., incident hospitalization counts during a flu season). Each week, a forecaster issues a forecast distribution *P*_*t*_ for *Y*_*t*_ using information available up to week *t* − 1. We denote the score of this forecast by *S*(*P*_*t*_, *y*_*t*_), where *S* is a proper score, such as the WIS or CRPS, and *y*_*t*_ is the observed count.

A common way to evaluate model forecast performance is using the arithmetic average of the scores (see e.g., [2, 8]). We refer to this score as the *prequential score* (PS) as in Dawid and Musio [11]:

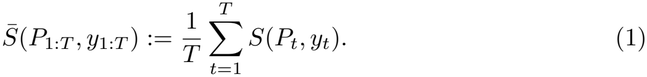

Here, we propose to instead evaluate forecasts using the *weighted prequential score* (WPS),

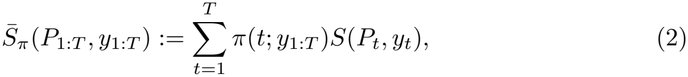

where *π*(*t*; *y*_1:*T*_) is the weight for time *t* depending on the observed sequence *y*_1:*T*_. Note that the PS is a special case where the weight distribution is uniform *π*_0_(*t*) ≡ 1*/T* .

#### 2.1.1 Weight construction using entropic tilting

For simplicity, we choose a binary rule for assigning weights to forecast-observation pairs, but our approach can be extended to more flexible weighting rules (see Supplementary Material). In particular, a set of *event weeks E* is assigned a higher weight relative to weeks in the *non-event week* set *E*^*c*^. Specifically, the user selects an emphasis ratio *r* ≥1 that determines how strongly the evaluation prioritizes accuracy during the event weeks relative to the rest of the season. Setting *r >* 1 emphasizes accuracy on weeks in *E* relative to the PS, while *r* = 1 recovers the PS.

Entropic tilting [25, 26] is demonstrated in Fig 2: the weights obtained under ET assign higher probability to the event weeks than the PS weights while remaining uniform *within E* and *E*^*c*^. More formally, each choice of *r* corresponds to a desired total weight

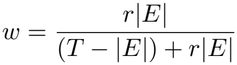

assigned to the event week set. ET allows us to construct the unique set of score weights that satisfies the constraint Σ_*t∈E*_ *π*(*t*; *y*_1:*T*_) = *w* while minimizing the Kullback–Leibler divergence from the PS weights. The WPS weights obtained by ET (derived in the Supplementary Material) lead to a WPS of the form:

**Fig 2.**
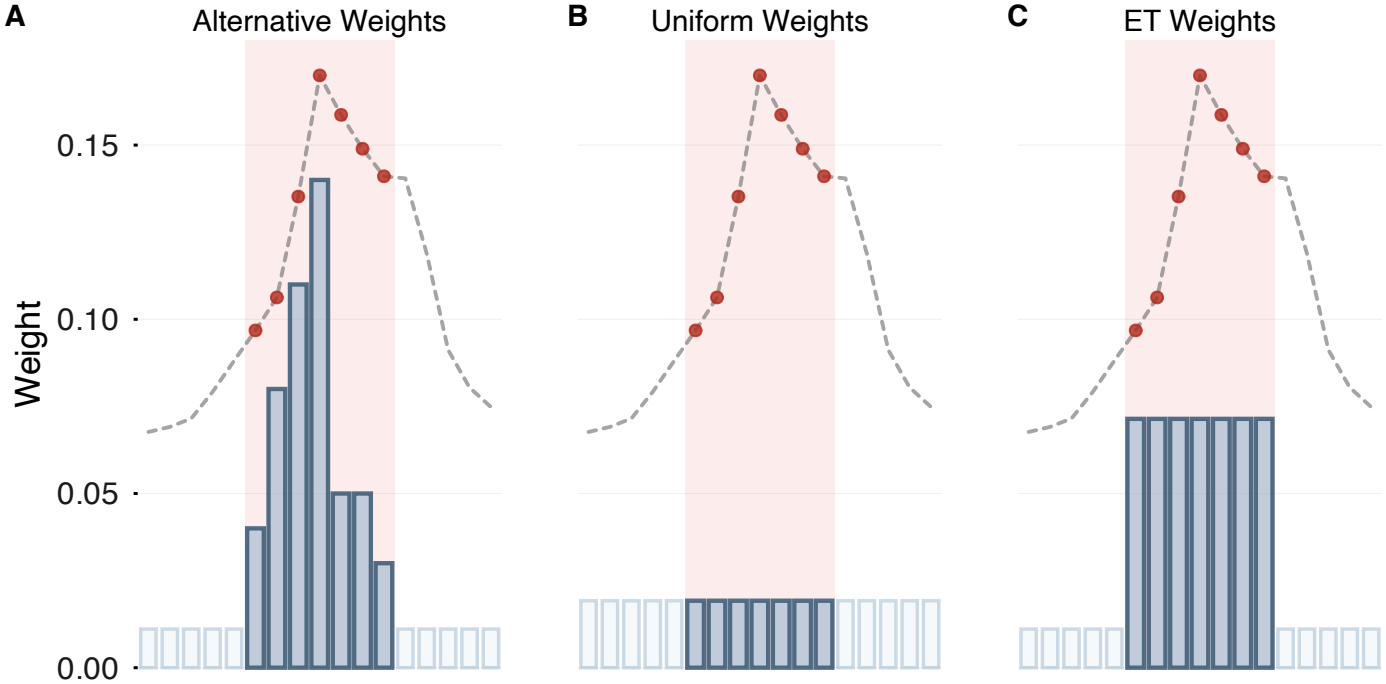
Illustration of weight distributions. The event weeks *E* = {*t* : |*t* − *t*_peak_| ≤ 3} are shaded in red, where *t*_peak_ denotes the week of the seasonal peak. **A**: Example alternative weight distribution that assigns total weight *w* to the shaded region but is not the ET weights. **B**: The uniform weight distribution used in the PS. **C**: The unique set of ET weights: uniform weighting is applied both inside and outside the shaded region so that weights are as close as possible to uniform while still assigning total weight *w* to the event weeks.

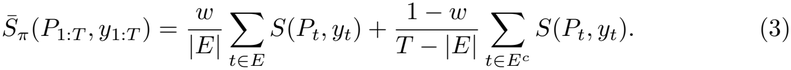

This formula can be understood as a two-stage average: first, compute the PS restricted to the event weeks *E* and the non-event weeks *E*^*c*^, then combine these scores as a weighted average with weights *w* and 1 − *w*.

### 2.2 Properties of the weighted prequential score

We quantify accuracy gains during event weeks and losses during non-event weeks when switching from the top-ranking PS model to the top-ranking WPS model. We then discuss the lack of propriety and the evaluation target induced under weighting.

#### 2.2.1 Improvement and penalty metrics

Let 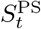be the score of the forecast for week *t* under the top-ranking PS model. For this model, the total scores over the event and non-event weeks, denoted ℰ^PS^ and *N*^PS^, respectively, are:

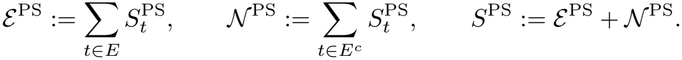

Similarly, we define 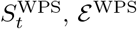, and *N*^WPS^ for the top-ranking WPS model. The *penalty* of choosing the top WPS model is the *increase* in the average non-event-week score relative to the average score of the top PS model:

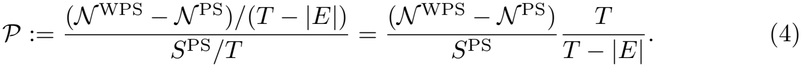

We divide by *S*^PS^ to allow for comparisons across different locations and seasons, since common scoring rules (e.g., WIS) depend on the observation scale. Conversely, we define the *improvement* as the average *decrease* in the event-week score relative to the average score of the top PS model:

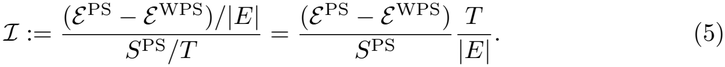

If the improvement ℐon event weeks exceeds the penalty *P* on non-event weeks, then the top WPS model may be preferable to the top PS model.

As the top WPS model achieves the lowest WPS across all models, we obtain the following inequality:

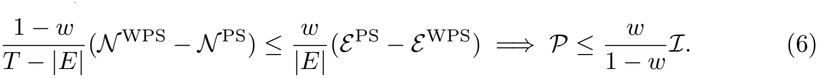

This inequality shows that the improvement is at least as large as the penalty whenever 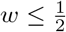 , since then *w/*(1− *w*) ≤ 1 and hence *P* ≤ ℐ : the average improvement on event weeks is at least as large as the average penalty on non-event weeks.

#### 2.2.2 Propriety and evaluation target

If the underlying scoring function *S* is (strictly) proper, then the PS aggregation leads to a (strictly) proper score for the joint distribution of *Y*_1:*T*_ [11]. Strict propriety implies that the expected score is uniquely minimized when *P*_*t*_ = *Q*_*t*_, where *Q*_*t*_ denotes the true probability distribution of *Y*_*t*_ given the past observations *y*_1:*t*−1_.

By definition, the weights obtained using ET depend on the observed outcomes *y*_1:*T*_. Consequently, the WPS is not proper for the joint distribution of *Y*_1:*T*_. Instead, the expected WPS is minimized by

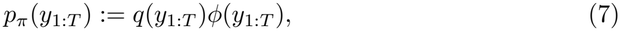

where *q*(*y*_1:*T*_) denotes the true probability mass function of *Y*_1:*T*_ and *ϕ* is a function that we derive in the Supplementary Material. Equation (7) implies that a forecaster who optimizes the WPS is incentivized to issue forecasts that assign higher probability to sequences *y*_1:*T*_ for which *ϕ* is large.

## 3 Application to FluSight model evaluation

FluSight [6, 8, 13] is a CDC-hosted forecasting challenge that collects real-time forecast submissions of incident influenza hospitalizations from multiple teams. At the end of a season, models are evaluated and ranked using their seasonal average WIS. We explore how weighted scoring changes these model rankings in three ways. First, we summarize how the top ranking model changes as the emphasis ratio *r* increases. Second, we show how contributions to the PS and WPS are distributed across a given season, and how upweighting errors during event weeks changes the rankings of the top five models.Finally, we end our analysis by examining what is gained or lost in terms of forecast accuracy by preferring the top WPS model.

For each analysis, we consider FluSight submissions for the 2023/2024, 2024/2025, and 2025/2026 seasons. We restrict to models with submissions for (i) at least 90% of the scorable forecast weeks and (ii) all event weeks under both the seasonal peak and high growth definitions given below; locations with fewer than 5 models satisfying these criteria were not considered in our analyses. We refer to a location-season-horizon combination as a *setting*.

### 3.1 Definition of higher-importance weeks

To demonstrate our framework, we propose two event-week definitions that emphasize high-growth weeks and weeks around the seasonal peak. These epidemic phases are especially difficult for models to capture, as illustrated by the consistent underprediction of high-growth weeks in the FluSight challenge [8]. Note that the WPS can be adapted to other event-week definitions (see Supplementary Material).

#### Seasonal peak event week definition

The event weeks for a given location are the set of weeks within a window of *τ* = 3 weeks around the seasonal peak. Mathematically, we have:

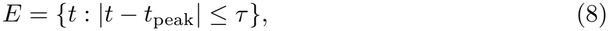

where *t*_peak_ denotes the week with the largest observed hospitalization count for a given location and season. This event-week definition is relevant to decisions where the timing and magnitude of the peak are central for preparedness (e.g., hospital staffing).

#### High-growth event week definition

The event weeks for a given location are the set of weeks with a large increase in incidence:

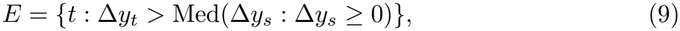

where Δ*y*_*t*_ := *y*_*t*_− *y*_*t*−1_ is the weekly change in incidence. *E* contains weeks with an increase in incidence greater than the median increase among all growth weeks (Δ*y*_*t*_≥ 0) at a given location.

We illustrate these event-week definitions in Fig 3 (red shaded area). Note that in these settings, the top-ranked models under the PS and WPS differ. In particular, observe that the top PS models produced precise forecasts on non-event weeks but undercovered on most of the event weeks; this suggests that their high rankings were primarily driven by strong performance on the non-event weeks. In contrast, the top-ranked model under the WPS had full coverage across the event weeks, though at the expense of performance on non-event weeks.

**Fig 3.**
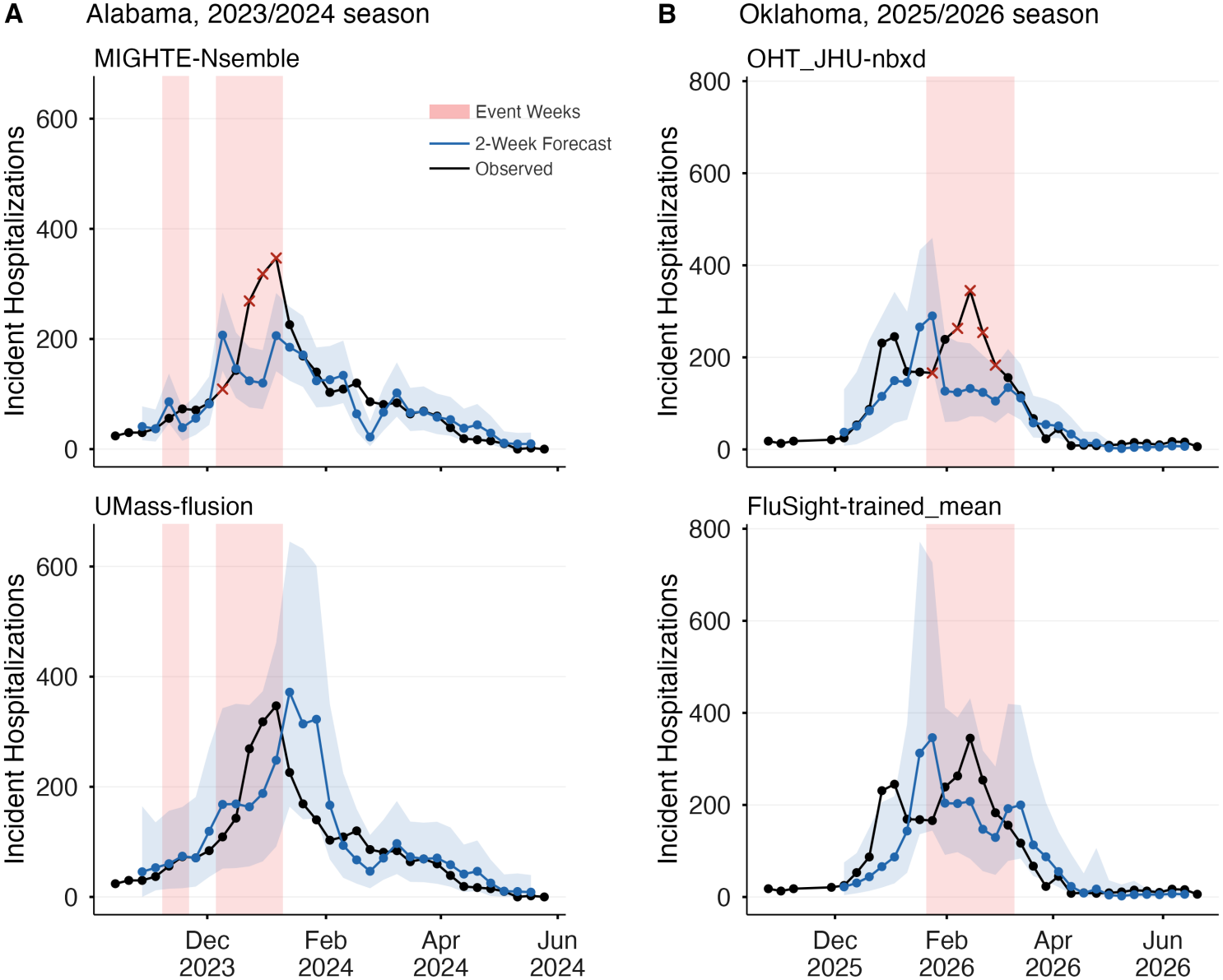
Examples of FluSight model ranking changes under the WPS. In both **A** and **B**, the top-ranking model under the PS is shown at the top, while the top-ranking model under the WPS is shown at the bottom. The black line denotes observed incident hospitalizations, and the 2-week horizon forecasts are denoted by blue lines. The blue shaded regions correspond to 90% prediction intervals. Red crosses indicate observations outside the prediction intervals. **A**: Forecasts issued for Alabama during the 2023/2024 influenza season. The WPS is defined using the high-growth event-week definition (9) with *r* = 2. **B**: Forecasts issued for Oklahoma during the 2025/2026 influenza season. The WPS is defined using the seasonal peak event-week definition (8) with *r* = 2.

### 3.2 Rank changes under weighted scoring

Switching from the PS to the WPS caused a change in model rankings that we explore for the US 2025/2026 season (Fig 4). Under the high growth priority, the WPS led to many model rank changes (9/34 models with a change≥ 3) and slightly reduced crowding. Conversely, the seasonal peak priority did not cause as many model rank changes (5/34 models with a change≥ 3). A potential reason for this stability is that the WIS is sensitive to observation scale, so the PS is primarily driven by performance during high-incidence periods.

**Fig 4.**
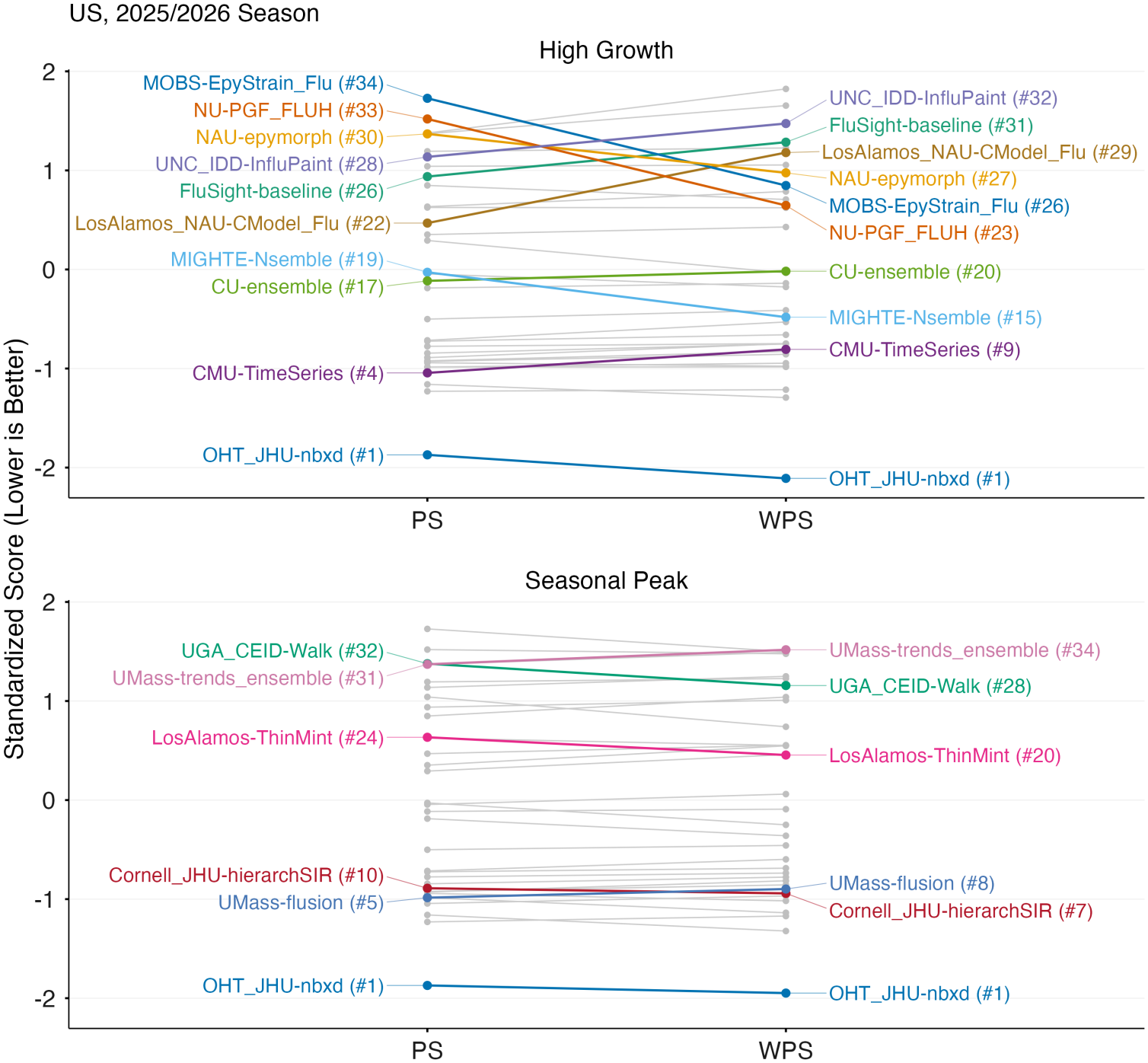
Slope plot comparing PS and WPS scores for the FluSight 2025/2026 season with *r* = 2. Models whose ranks changed by more than two positions are labeled, with ranks shown in parentheses next to the model names. Within each score-priority setting, scores are standardized using the corresponding mean and standard deviation. As with the WIS, lower standardized scores indicate better performance.

We now explore how top-model rankings change under weighted scoring. Recall that *r* corresponds to the ratio of the event and non-event week weights, meaning that an event week receives *r* times the weight of a non-event week in the WPS. In Table 1, we see that the percentage of locations where the top PS and WPS models agree for all *r* is between 37% and 68% across seasons and horizons. In contrast, the percentage of locations where the top models differed was between 10% and 26% for *r* ≤2, and between 17% and 43% for *r*≤ 4. Thus, the top-ranking model is robust to event-week emphasis in some settings, whereas in others even a twofold emphasis changes the preferred model.

**Table 1.** Summary of top model rank changes across locations by season and horizon *h*.

| Event | Season | *h*<sup>a</sup> | *n*<sup>b</sup> | Same all *r* %<sup>c</sup> | Diff. *r* ≤ 2 %<sup>d</sup> | Diff. *r* ≤ 4 %<sup>e</sup> |
|---|---|---|---|---|---|---|
| Peak | 2023/2024 | 2 | 48 | 46% | 19% | 35% |
|  |  | 3 | 50 | 52% | 14% | 38% |
|  | 2024/2025 | 2 | 52 | 40% | 12% | 29% |
|  |  | 3 | 48 | 38% | 21% | 40% |
|  | 2025/2026 | 2 | 51 | 55% | 26% | 37% |
|  |  | 3 | 38 | 68% | 16% | 26% |
| Growth | 2023/2024 | 2 | 48 | 44% | 21% | 27% |
|  |  | 3 | 50 | 66% | 12% | 20% |
|  | 2024/2025 | 2 | 52 | 67% | 10% | 17% |
|  |  | 3 | 48 | 67% | 13% | 19% |
|  | 2025/2026 | 2 | 51 | 37% | 22% | 43% |
|  |  | 3 | 38 | 53% | 13% | 26% |

<sup>a</sup> Forecast horizon length in weeks.
<sup>b</sup> Number of eligible locations used to construct the estimated CDFs in Fig 7.
<sup>c</sup> Percentage of locations where the top-ranked PS and WPS model are equal for all *r*.
<sup>d</sup> Percentage of locations where the top-ranked PS and WPS models disagree when *r* ≤ 2.
<sup>e</sup> Percentage of locations where the top-ranked PS and WPS models disagree when *r* ≤ 4.

Fig 5 shows the contribution of weekly scores to the PS and WPS across the 2025/2026 season for the top five models. We see that the rankings differ under the WPS, demonstrating how the WPS prioritizes accuracy during the period of the season emphasized by the user. As an example, consider the `NAU-vulPES` model (yellow line). It was ranked 2nd under the PS since its non-event week contributions to the PS were typically smaller than those of the other models. However, its PS contributions during the event weeks tended to be larger, and consequently it fell in ranking to 4th under the seasonal peak WPS, and 3rd under the high growth WPS.

**Fig 5.**
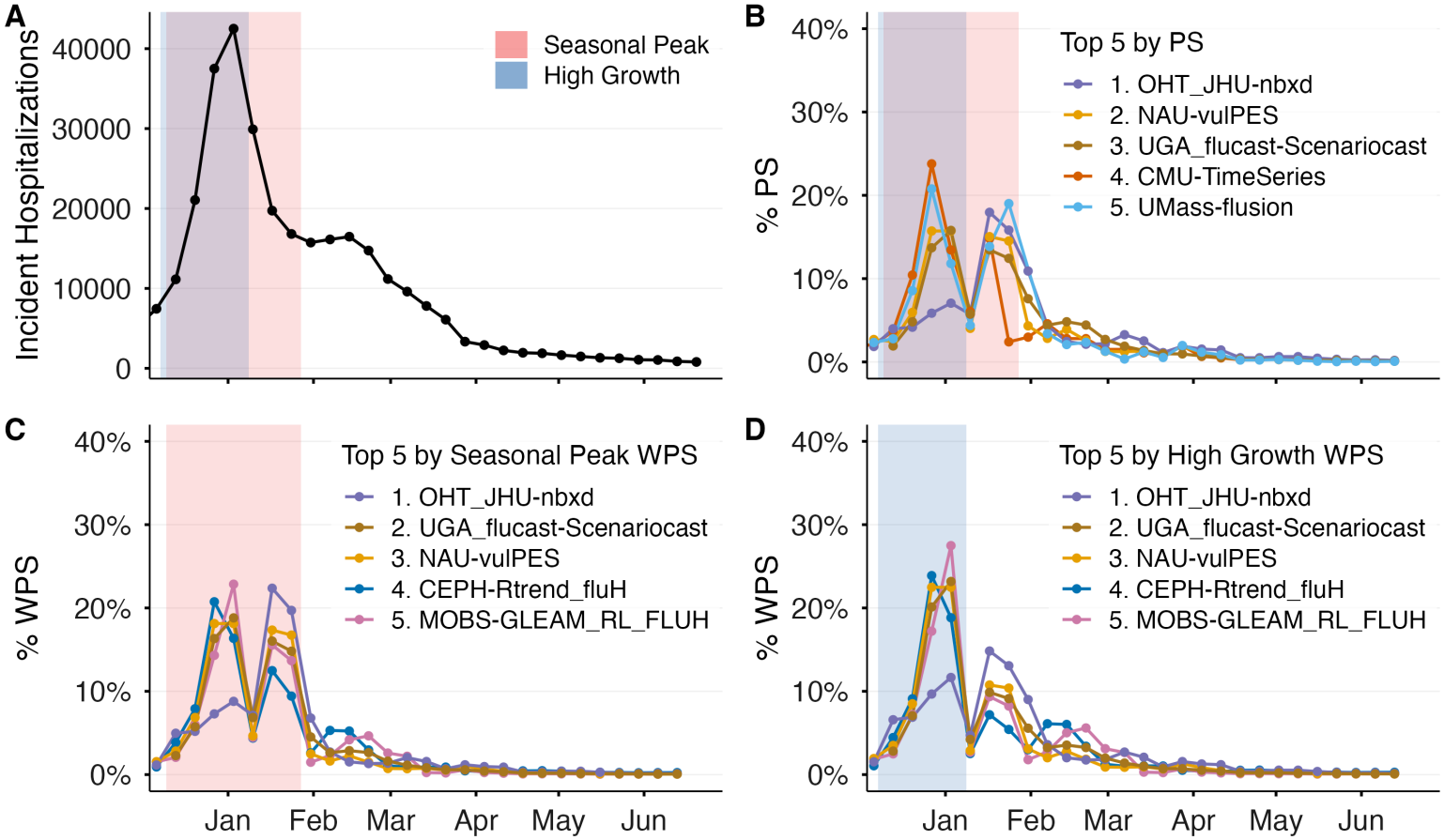
Percentage contribution of weekly scores to the PS and WPS across the U.S. 2025/2026 season for the top 5 ranking models. In both cases, *r* = 2 and the forecast horizon is two weeks. The blue shaded region corresponds to the high-growth event weeks, and the red shaded region corresponds to the seasonal peak event weeks. **A**: Observed incident hospitalization count. **B**: Contribution of weekly scores to the PS for the top 5 ranking models. **C**: Contribution of weekly scores to the WPS for the top 5 models under the seasonal peak event week definition. **D**: Contribution of weekly scores to the WPS for the top 5 models under the high-growth event week definition.

### 3.3 Event week score improvements under the WPS

We analyze the improvement ℐ in WIS during event weeks and the penalty *P* during non-event weeks under the WPS (see (4) and (5)). In Fig 6, we display ℐ and *P* across all seasons, horizons (2 and 3 weeks), and locations for the peak event-week definition, varying the total event-week weight *w* ∈ {0.3, 0.5, 0.9}. In agreement with our theoretical result (6), we see that 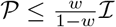, as all points lie in the gray shaded region.For *w* ∈ {0.3, 0.5}, nearly all observations are above the *P* = ℐ line. In this case,switching from the top-ranked PS model to the top-ranked WPS model leads to a higher event-week improvement than non-event-week penalty. Conversely, for extreme weighting *w* = 0.9, many observations have a higher penalty than improvement, indicating that the WPS model comes with a substantial trade-off. Note that an improvement of 0.5 means that the average event-week decrease in WIS is 50% of the top PS model score.

**Fig 6.**
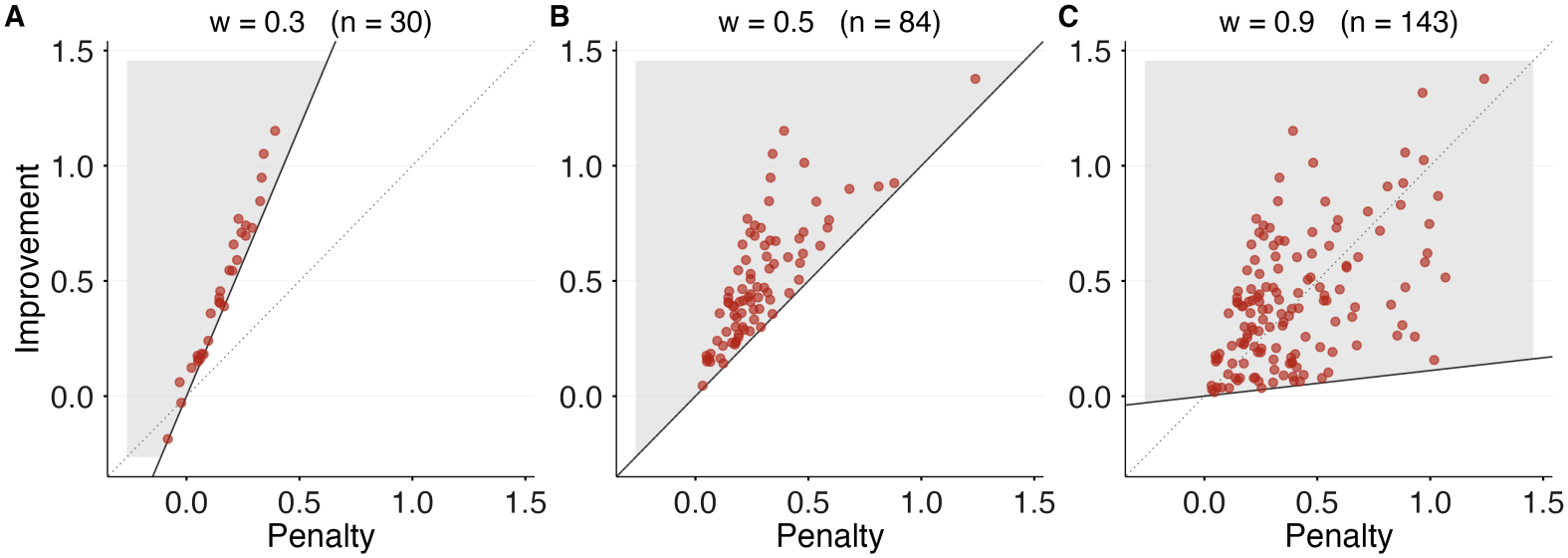
Improvement and penalty trade-off across different WPS weight values. Each point corresponds to a location, season, and horizon setting where the top-ranked PS and WPS models differed. The dotted line corresponds to the *P* = ℐ line. The region satisfying the theoretical bound in (6) is shaded in gray. **A**: The PS and WPS frequently agree on the top model since *w*≈ |*E*| */T*. However, when they disagree, the improvement is substantially greater than the penalty. **B**: At *w* = 0.5, the bound guarantees that improvement is at least as large as penalty. **C**: Due to the extreme weighting, the top-ranked WPS and PS models differ frequently, and the penalty is larger than the improvement across many settings.

## Discussion

By highlighting differences in model performance that may be obscured by averaging across weeks within a season or epidemic, the WPS provides a complementary diagnostic to proper scoring rules commonly used in forecast evaluation. Importantly, it requires only a definition of event weeks and the emphasis ratio, *r*, making it particularly useful when an explicit utility function is difficult to specify. We demonstrated on the CDC FluSight challenge that the WPS is a flexible method to underscore priority periods in model evaluation, and that under suitable weights it selects models whose improvements during event weeks exceed losses during non-event weeks.

The WPS is closely related to existing scoring rules that connect the quality of a forecast to its value for public health decision making. Notably, Gerding et al. [15] derive a proper scoring rule that evaluates forecasts based on their ability to effectively allocate limited medical resources; Marshall et al. [16] propose a modification of WIS that normalizes and caps the interval score according to a utility-based error tolerance. These approaches provide a valuable framework for connecting a given scoring rule to an explicit utility function. The WPS takes a complementary approach by changing how an unmodified score is aggregated over a season. The benefit of this approach is that neither a loss function nor a utility threshold needs to be defined explicitly, as these may be difficult to define or elicit. Both strategies are nonetheless closely related, since a utility-based scoring function implicitly emphasizes a particular set of weeks. For example, the allocation score is largest when incidence is close to the amount of available resources, which in the application of Gerding et al. occurred during the growth and decline periods of the epidemic curve [15]. Another alternative scoring rule proposed by Bosse et al. involves scoring epidemic forecasts on a log-transformed scale [27]. This approach results in a proper scoring rule which mitigates the influence of high-incidence weeks or highly populated locations on the aggregate seasonal score. At the expense of propriety, the WPS can achieve a similar effect for an appropriately defined set of event weeks, additionally allowing more control over the amount of emphasis (e.g., giving more weight to less populated locations while still emphasizing performance on high incidence weeks).

The main limitation of the WPS is that, when weights depend on observed outcomes, it is not proper for the true joint data-generating distribution. Instead, the WPS incentivizes forecasters to assign higher probability to outcome sequences that receive larger weight under the WPS. This issue is closely related to the *forecaster’s dilemma* identified by Lerch et al. [17], who showed that when evaluation is restricted to extreme observations, forecasters do not need to be skillful with respect to the true data-generating distribution. Consequently, we recommend using the WPS as a retrospective performance diagnostic rather than as a replacement for PS-based evaluation. Nonetheless, we show in the Supplementary Material that the WPS can be *properized* using a Bayes-act construction [28]. This properized WPS can be approximated using Monte Carlo integration when simulations from the forecast distribution are available. However, this approach cannot currently be applied to submissions from forecasting hubs such as FluSight, as they collect forecast quantiles instead of trajectories. Nonetheless, the increasing availability of trajectory-based forecast submissions could make properized versions of the WPS feasible in future applications [29].

Forecast accuracy has been shown to degrade during periods of rapid epidemic change [8], and, more generally, to vary across epidemic phases [30]. Yet these differences may be obscured when models are evaluated using seasonal average scores, particularly when important event weeks represent only a small fraction of the evaluation period. By allowing users to explicitly prioritize the weeks most important to their decisions, the WPS provides a flexible framework that makes the temporal priorities underlying aggregate evaluation explicit and better connects predictive performance to its downstream value for decision making. Consequently, we recommend using the WPS with moderate weighting alongside proper scoring rules to support model selection that better reflects public-health priorities.

## Data Availability

The code to reproduce all graphs and results in this paper is available at https://github.com/ACCIDDA/weighted-prequential-score-code, for which a DOI will be provided after revision.

## Acknowledgments

This project was made possible by cooperative agreement CDC-RFA-FT-23-0069 from the CDC’s Center for Forecasting and Outbreak Analytics. Its contents are solely the responsibility of the authors and do not necessarily represent the official views of the Centers for Disease Control and Prevention. The authors thank Paul Zivich for valuable advice, Tijs Alleman for manuscript comments and Nicholas Reich for pointing out the link to the forecaster’s dilemma.

## Supplementary Material

## 4 Sensitivity analysis of model rankings to WPS

We define *r*_crit_ as the smallest emphasis ratio *r* for which the top-ranking models differ under the PS and WPS. When *r*_crit_ =∞ , the rankings are robust to priority weighting, as the top-model rankings under the PS and WPS agree for any *r*. Conversely, lower *r*_crit_ values mean that the top-model ranking is less robust to changes in the weights.For each season and horizon (2 and 3 weeks), we compute 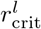 for each location *l* to form an estimate of Pr(*r*_crit_ ≤ *r*). In Fig 7, we show the estimated probability Pr(*r*_crit_ ≤ *r*) across locations as *r* increases.

**Fig 7.**
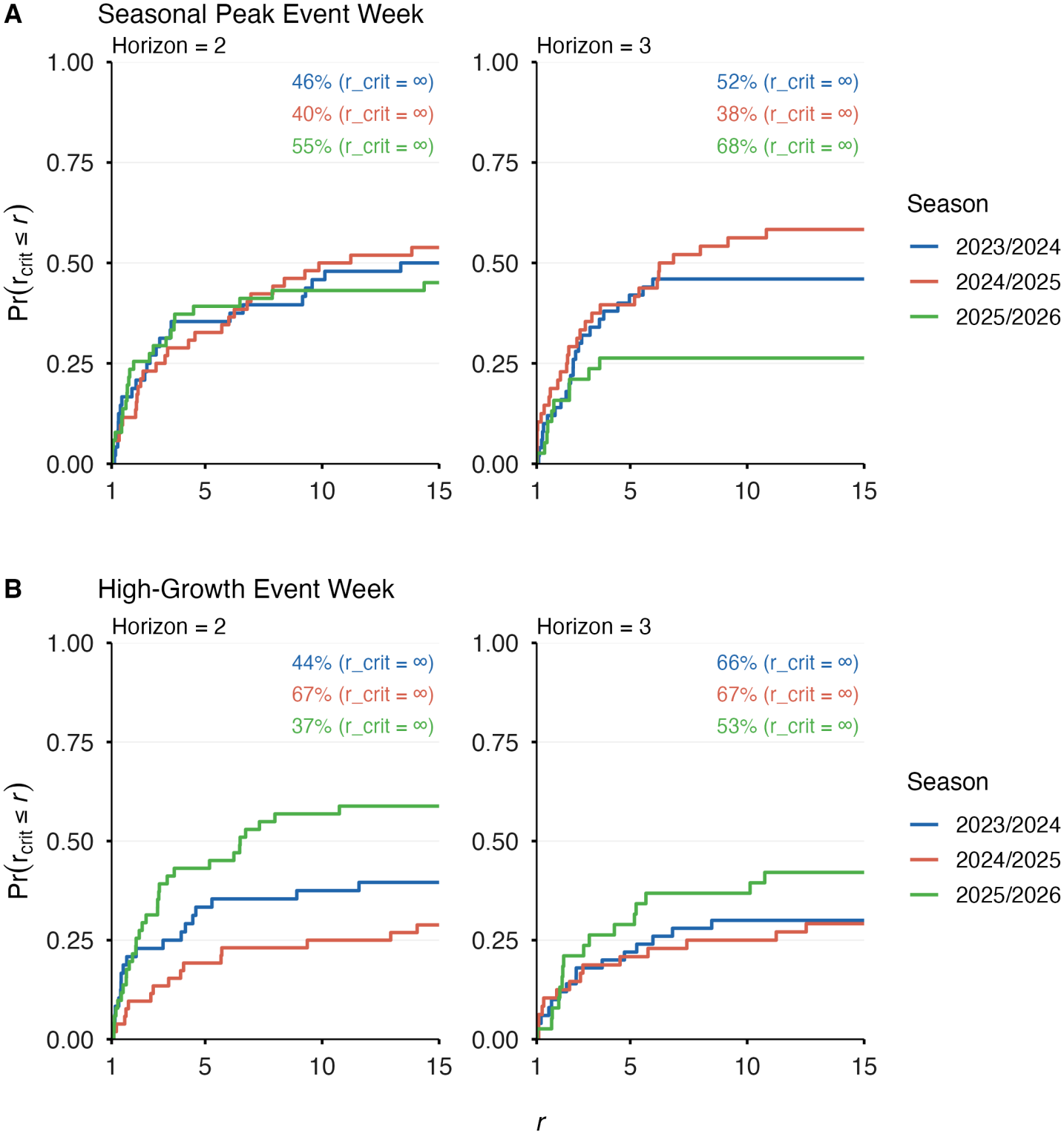
Analysis of the stability of model rankings between the WPS and PS. We show Pr(*r*_crit_ ≤ *r*) as a function of the emphasis ratio *r*, along with the percentage of locations with *r*_crit_ = ∞, for the **A**: seasonal peak weight function (8) and **B**: high-growth weight function (9).

## 5 Entropic Tilting

The description of entropic tilting (ET) given here suffices to describe the WPS construction; a more general description can be found in [25].

### 5.1 WPS Construction

Let Δ denote the set of probability distributions defined on {1, … , *T*} with full support. Suppose that we would like to obtain a distribution *µ* ∈ Δ that satisfies

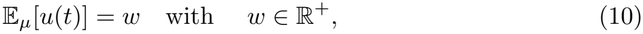

where *u* : {1, … , *T}*} →ℝ^+^ is a non-negative function. Note that many distributions satisfy (10). Now suppose that we would like to obtain *µ* that satisfies (10) *and* remains as ‘close’ as possible to a prior distribution *π*_0_ ∈ Δ. Formally, let

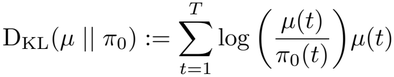

denote the KL-divergence from *µ* to *π*_0_. We seek a distribution *π* satisfying

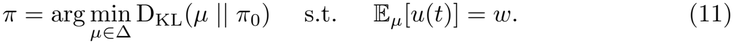

The (unique) solution to (11) can be found using Lagrange multipliers under suitable conditions. In particular, it can be shown that *π*(*t*) ∝ *π*_0_(*t*) exp(*λu*(*t*)) is an exponentially tilted version of *π*_0_, where *λ* is the Lagrange multiplier. When 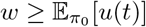, we have *λ* ≥ 0, and the equality constraint can be relaxed to E_*µ*_[*u*(*t*)] ≥ *w* without changing the solution (see [26]). For the construction of the WPS, we choose *π*_0_(*t*) ≡ 1*/T* and therefore obtain the following expression for the ET weights:

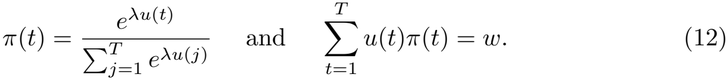

Note that *λ* determines how concentrated *π* is on weeks where *u* is large. When *u* is an indicator function, *λ* admits a closed-form expression (see below). However, in general *λ* must be computed numerically using a root-finding algorithm.

### 5.2 Indicator Weight Functions

Suppose *u*(·; *y*_1:*T*_) : {1, … , *T*} → {0, 1} is an indicator weight function. Recall *E* = {*t* : *u*(*t*; *y*_1:*T*_) = 1} denotes the set of event weeks. Then the moment constraint (10) reduces to

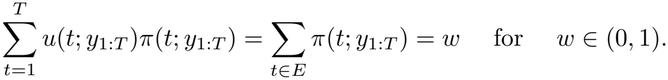

Therefore *w* controls the total probability mass assigned to the event weeks *E*. Under the prior distribution *π*_0_, this mass equals Σ _*t*∈*E*_ *π*_0_(*t*) = |*E*|*/T*. Consequently, *π* = *π*_0_ is feasible if *w* = |*E*|*/T* (and in this case is also the minimizer of D_KL_(· || *π*_0_)), whereas if *w >* |*E*|*/T* then the ET solution upweights *E* relative to *π*_0_. More generally, for *w* ∈ (0, 1), it is straightforward to check that the tilting parameter is *λ* = log(*r*), where we recall 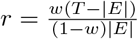 , leading to the following expression for the ET weights:

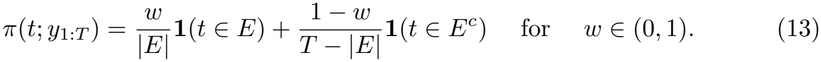

(Note that *E* is implicitly a function of *y*_1:*T*_). Observe that *π*(*t*; *y*_1:*T*_) is piecewise constant in (13), meaning that *π* is a mixture of uniform distributions with mixture weight *w*. Such a solution is not surprising since *π* must assign *w* weight to weeks in *E and* minimize D_KL_(· || *π*_0_). Plugging (13) into (2) gives us the weighted score (seeFig 2).

## 6 WPS Target and Properization

### 6.1 Setup

Let *Q*_1:*T*_ denote the true joint distribution of the observations *y*_1:*T*_ and *P*_1:*T*_ be a corresponding joint forecast distribution; at times, we will omit the subscript and write *Q* and *P* for brevity. Recall that the WPS is given by

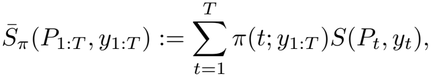

where we assume *S* is a proper scoring rule and *P*_*t*_ the forecasted distribution of *Y*_*t*_ conditional on observing *Y*_1:*t*−1_ = *y*_1:*t*−1_:

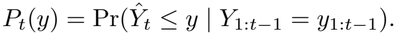

By definition, the target distribution of the WPS is the joint forecast distribution 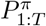 that minimizes the expected score:

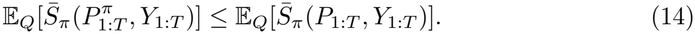

### 6.2 Definition of ϕ(y_1:*T*_)

Let 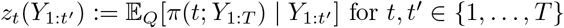 and *c*_1_ = E_*Q*_[*π*(1; *Y*_1:*T*_)]. The function *ϕ*(*y*_1:*T*_) is defined as

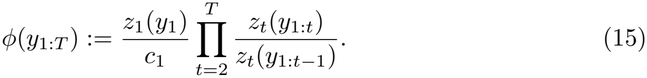

We claim that the PMF of the minimizer 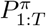 is given by *p*_*π*_(*y*_1:*T*_) := *q*(*y*_1:*T*_)*ϕ*(*y*_1:*T*_), where *q* denotes the PMF of *Q*. Indeed, observe that 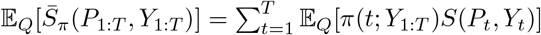. Now write for *t* ∈ {1, … , *T*} using the tower property

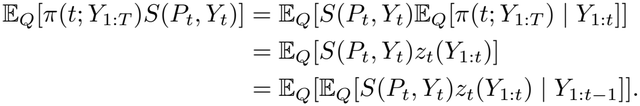

Focusing on the inner expectation, we have

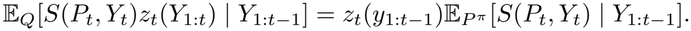

Since *S* is proper, it follows that E_*P*_ ^*π*^ [*S*(*P*_*t*_, *Y*_*t*_)] is minimized by choosing 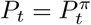 for *t* = 1, … , *T*. Hence, 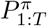minimizes 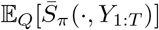, establishing (14).

The tilt *ϕ*(*y*_1:*T*_) given in (15) implies that a forecaster optimizing the WPS is incentivized to issue forecasts that assign a higher probability to sequences *y*_1:*T*_ where we tend to have

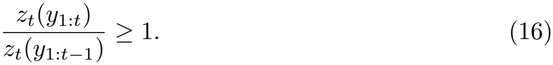

For example, suppose *u*(*t*; *y*_1:*t*_) is an indicator weight function depending only on *y*_1:*t*_ and that *w >* |*E*| */T*. Then the inequality (16) holds “on average” for high weight weeks in that one can show

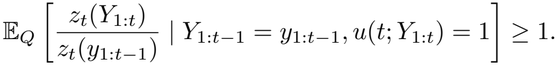

### 6.3 Properization

Let *P* be a forecast distribution with joint PMF *p*(*y*_1:*T*_). Let *T* be a map that assigns to *P* a *Bayes act* under 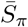 in the sense that *T* satisfies

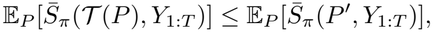

for any other forecast distribution *P* ^′^. In particular, the results from the previous section show that the mapped forecast distribution *T* (*P*) has PMF 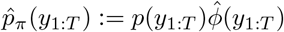, where

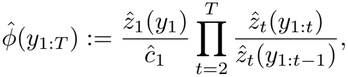

with 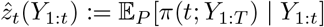and *ĉ*_1_ := E_*P*_ [*π*(1; *Y*_1:*T*_)]. Note that 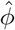 is identical to (15) except that expectations are taken with respect to the forecast distribution *P*. As discussed in [28], by identifying *T* we are able to obtain a *properized* WPS 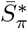 that is proper for the true joint data distribution *Q*:

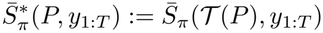

Indeed, this fact follows immediately from the results of the previous section, where we showed that (14) holds with *T* (*Q*) = *P* ^*π*^. By definition, the properized WPS is proper for the true data-generating distribution: the joint distribution of *Y*_1:*T*_ .

Since 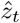is defined as an expectation under the *forecast distribution*, one can approximate it numerically via Monte Carlo provided forward simulation of *Y*_*t*+1_, … , *Y*_*T*_ is possible under *P*. We believe that this is the case for many current forecasting models and could be done a posteriori in forecast hubs that recorded forecast trajectories (but not from quantiles submissions). However, forecasting hubs such as FluSight currently only require that teams submit quantiles under *P* instead of simulations.

